# Wei Ren Anti-Wart Formula combined with photodynamic therapy for Condyloma acuminate: a double-blind, randomized, parallel-group trial

**DOI:** 10.1101/2024.01.10.24301098

**Authors:** ShiYan Yang, Liang Hua, Dongjie Guo, Yifei Wang, Xuqiang Wei, Fulun Li

## Abstract

**Introduction:** Condyloma acuminate (CA) is a chronic disease with a high rate of recurrence which has a detrimental impact on patients’ physical and mental health, as well as their quality of life and socioeconomic advancement. Photodynamic therapy is a less invasive and selective intervention for CA, but its safety and high price limit its utilization. Wei Ren Anti-Wart Formula (WRAWF), is a promising Chinese medicine for CA, however, the evidence of its effectiveness and safety is scarcely. This trial aimed to evaluate the clinical effectiveness, safety, and recurrence management of WRAWF combined with photodynamic in the CA patients.

**Methods and analysis:** A double-blind, randomized, parallel-group trial was designed. Participants will recruit in the Yue Yang Traditional Chinese Medicine Hospital, Shanghai University of Traditional Chinese Medicine from May 1, 2024, to December 31, 2025. The sample size is 148 CA patients (74 in each group), will randomly divide into the trial and control groups. followed up at weeks 6, 10, and 14 after the 2-week treatment period. The recurrence rate will be set as the primary outcome. The visual analogue scale (VAS) scores of pain, the rate of infection, superficial scarring, and the type, severity, and incidence of adverse events will be carefully documented at 0,1,2,6,10,14 weeks.

**Ethics and dissemination:** This trial has approved by the Ethics Committee of Yueyang Hospital of Integrated Traditional Chinese and Western Medicine affiliated to Shanghai University of Traditional Chinese Medicine (No. 2023-023). Meanwhile, informed consent was provided by all patients conducted in accordance with the Declaration of Helsinki.

**Trial registration number:** ChiCTR2300071609; www.chictr.org.cn;Registered on April19, 2023(first version).

## BACKGROUND

Condyloma acuminate (CA), also known as anogenital warts, is a sexually transmitted disease caused by human papillomavirus (HPV) infection that is primarily a warty, proliferative lesion of the skin and mucous membranes. It typically affects the skin and mucous membranes of the vaginal, anal, and perianal areas, and yet it can also affect the inguinal and perineal areas. It has a very high recurrence rate, with a large sample showing a recurrence rate of (47-163)/100,000 in mele and (23-110)/100,000 in female.[1–4] As a result, CA is prone to recurrence, with the maximum recurrence rate occurring within 3 months, and then progressively declining as time goes on. Therefore, it requires long-term, frequent treatment. Additionally, there is a significant influence on the patient’s everyday life and mental health.

Photodynamic therapy (PDT) is a topical application of the photosensitizer 5-aminoketovaleric acid (ALA), followed by topical light treatment with a semiconductor laser or light emitting diode (LED), usually using red light (630-635 nm). It is indicated for the removal of smaller warts and basal treatment after physical therapy has removed larger warts. The treatment was performed with a 10%-20% solution of 5-ALA applied to the lesion, extending to 1 cm around the wart, covering the subclinical area of infection around the wart as much as possible. For external genital warts, 3-4 sessions are recommended as a course of treatment for initial cases and 4-6 sessions for recurrent and recalcitrant cases.[5] The advantage is that it is non-destructive compared to other therapies such as laser and freezing, can be repeated, and is less likely to cause tissue defects and dysfunction. However, its high treatment price limits its clinical application.

Wei Ren Anti-Wart Formula (WRAWF), a Chinese herbal prescription, is a compound preparation with eight ingredients (Clematis chinensis Osbeck, Coicis Semen, Salvia miltiorrhiza Bunge, Astragali Radix, Atractylodes macrocephala Koidz,Hedyotidis Herba,Portulaca and Curcumae Rhizoma), achieving clinical efficacy in the external treatment of CA. It has the advantages of simple use, low treatment cost and few side effects. External use can act on the wart body.

Modern studies have shown that all the extracts of clematichinenoside have significant anti-inflammatory, analgesic, and antispasmodic effects.[6] The triglyceride component of Coix lacryma has anti-tumor effects, and its analgesic and anti-inflammatory effects have also been confirmed.[7,8] The mechanism of action of Salvia miltiorrhiza may be to suppress the inflammatory response of endothelial cells by down-regulating the expression of CD40 signals.[9] Astragalus polysaccharides have protective effects against oxidative stress injury and immune damage, and they promote immune function in traumatic tissues and facilitate wound repair.[10–12] Curcumin, the main component of Curcuma longa, has antitumor and antioxidant effects, and its ability to effectively scavenge free radicals leads to a reduction in the possibility of cellular carcinogenesis, and its mechanism of action meets the requirements of modern medicine needed for anti-photodamage drugs.[13] Therefore, the combination of the above-mentioned herbs and photodynamic therapy can not only play a role in reducing the recurrence rate of CA but also synergistically reduce the side effects of treatment and improve the regional immune function with photodynamic therapy.

Therefore, this trial adopts a A double-blind, randomized, parallel-group design to This trial evaluate the clinical effectiveness, safety, and recurrence management of the WRAWF combined with photodynamic in the CA patients objectively. The result will obtain high-level evidence-based medical evidence of Chinese medicine in the treatment of CA. This protocol will be reported in line with the SPIRIT checklist.

## METHODS

### Design

This is a double-blind, randomized, parallel-group clinical trial. The aim is to elucidate the clinical efficacy, safety, and recurrence of WRAWF combined with photodynamic therapy. The results of the study will establish the clinical standard for sequential treatment of CA with Chinese and Western medicine. The trial protocol (version 1.0, July 3, 2022) is being written and enrollment in the trial has not yet begun. Recruitment is expected to begin on May 1, 2024, and end on December 31, 2025.

Participants will recruit from the outpatients of Yueyang Hospital of Integrative Medicine, affiliated with Shanghai University of Traditional Chinese Medicine. A total of 148 patients will be enrolled in this clinical trial.

The study consists of five phases: screening/enrollment, allocation, treatment/intervention, end of intervention, and follow-up. In the enrollment process, participants will be recruited via a Condyloma acuminate clinic for physical examination and eligibility assessment. The time between the assessment and the intervention should not be longer than one week (week 0). If more than a week has passed since the assessment, assessment of the participant will be repeated before the intervention. If eligible, the participants will be requested to sign the written informed consent regarding participation in the trial (procedures, risks, options for dropping out), the use of laboratory data, and collection, storage, and use of biological specimens. Details of the informed consent will be explained by study investigators or medical staff members with adequate training. Once informed consent form is signed and dated by the participant, a participant identification (PID) number will be assigned to facilitate participant identification of the study.

For the random distribution of WRAWF groups and placebo group, randomisation will be performed by drawing lots, using the research randomiser program (https://www.randomizer.org/). The allocation sequence will be made by a different researcher,who will not be involved in the procedures.

The participants will be randomly allocated to the WRAWF group or placebo group. PIDs will be grouped by research randomiser program (https://www.randomizer.org/). We will treat the participants in the WRAWF group with WRAWF and PDT, those in the placebo group with WRAWF placebo and PDT (Fig 1). The participants will receive systemic therapy as per the judgment of their treating physicians. We will record all changes in symptoms, prescriptions, relevant scores, and macroscopic characteristics (based on photographic evidence), as well as any adverse events.

**Figure 1.** Flow diagram showing progress through the study.

The trial protocol was approved by the Ethics Committee of Shanghai Yueyang Hospital of Integrative Medicine and registered in the Clinical Trials Registry. Registration number: 2023-023

### Eligibility criteria

Inclusion criteria

a. clinical diagnosis of CA, between 18 and 65 years of age; both sexes.
b. warts located in the perianal area, distal urethra, and external genitalia.
c. who voluntarily participated in this study and signed the informed consent form.

Exclusion criteria

a. Those who have undergone ALA-PDT treatment within 12 weeks and those who apply any other treatment that affects the study.
b. Those with a combination of other conditions that may affect the evaluation of efficacy, such as other photo dermatological conditions, combination of severe localized infections.
c. those who have taken phototoxic or photosensitizing drugs within 12 weeks
d. clinical and/or pathological evidence of tumor invasion to other organs or tissues.
e. Severely immunocompromised individuals.
f. Scarred individuals.
g. Those with known cutaneous photosensitivity, porphyria, or allergy to ALA, light, or lidocaine
h. Those with severe medical, psychological, and psychiatric illnesses, infectious diseases, or pregnant or lactating women.
i. Any other reason deemed by the investigator to be inappropriate for participation in this study.

### Intervention

#### Trial group intervention

On day 0, day 7, and day 14 of the trial, we gave WRAWF topical application + Aminoglutaric Acid Hydrochloride topical dispersion + He-Ne laser irradiation once respectively.

Specifically: the test group will get 1 packet of WRAWF granules per dose, put all the granules of each packet into the same container, pour in about 5 ml of water for injection, stir until the granules are dissolved, and apply them externally for 30 minutes. Later Aminoketovaleric Acid Hydrochloride topical application is dissolved in water for injection before use (118mg/bottle add 0.5ml), prepared into a solution with a concentration of 20%. After cleaning the affected area and keeping it dry, put the 20% solution of amino glutarate hydrochloride on the cotton ball and cover the surface of the wart, repeat the solution on the cotton ball every 30 minutes or so, and keep dressing on the affected area for not less than 3 hours. The entire procedure should be done in a light-proof environment, and the affected area should be protected from direct light after the application of the medication. Subsequently, a helium-neon laser is used, with an output wavelength of 632.8 nm and laser energy of 100-150 J/cm, and the treatment spot completely covers the lesion.

#### Control group intervention

Placebo topical application + Aminoketovaleric acid hydrochloride topical dispersion + He-Ne laser irradiation

Specifically: Using placebo topical, placebo herbal granules were composed of a matrix, color correcting agent, flavoring agent, and a solvent selected according to the ingredient ratio, made by mixing, drying, crushing, sieving, granulation, and whole granules; the appearance, odor, and color were the same as the test drug. The remaining treatment method was the same as the test group.

#### Outcome measures

By employing deterministic, A double-blind, randomized, parallel-group trial, we aim to furnish precise directives for the secure management of patients diagnosed with Condyloma acuminate. Data derived from the envisaged outcome metrics will be amassed for subsequent investigations encompassing larger cohorts. Parameters such as recruitment and retention rates, the percentage of patients failing to meet eligibility criteria, and patients’ inclination towards random assignment will be computed. Additionally, assessments will be conducted on compliance levels, qualitative feedback, and compliance rates. Economical data pertaining to the utilization of medical resources and health-related quality of life will be gathered and subjected to comprehensive analysis. Safety outcome measures will be presented in compliance with the stipulations outlined for a clinical trial designed to scrutinize a pharmaceutical regimen.

#### Primary outcome

The main effect indicator was the time to recurrence: the date when a new wart was found at the treatment site from the first treatment.

#### Secondary outcome

The secondary effect index is the efficacy evaluation criteria - (1) cured: all warts were cleared after treatment and no new warts appeared; (2) significantly effective: the lesion area was reduced by ≥ 90% after treatment; (3) effective: the lesion area was reduced by 60% to 89% after treatment; (4) ineffective: the above criteria were not met. Total effective rate = (cured + effective + effective) / total number of cases × 100%. The incidence of pain visual analogue scale (VAS) score, infection, and superficial scar was also recorded during the treatment period, and the patients were scored for pain, infection, and scar. VAS was used for pain assessment; the Vancouver Scar Scale (VVS) was used to count and score the incidence and nature of scarring at the treatment site that appeared late after the patients underwent treatment.(Table 1, 2) At the same time, the type, degree, and incidence of adverse events will be recorded.

**Tables1.**
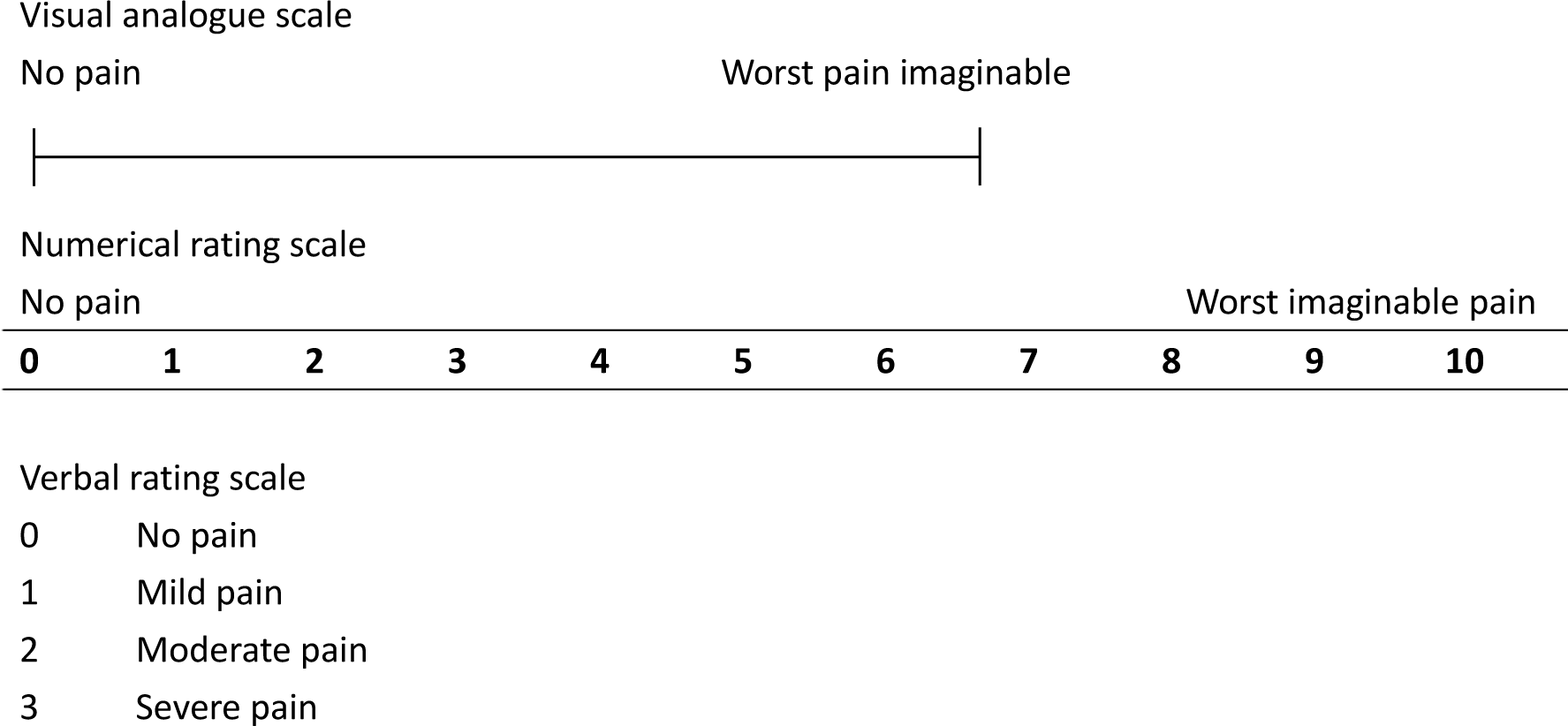
Visual analogue scale (VAS)

**Tables2.**
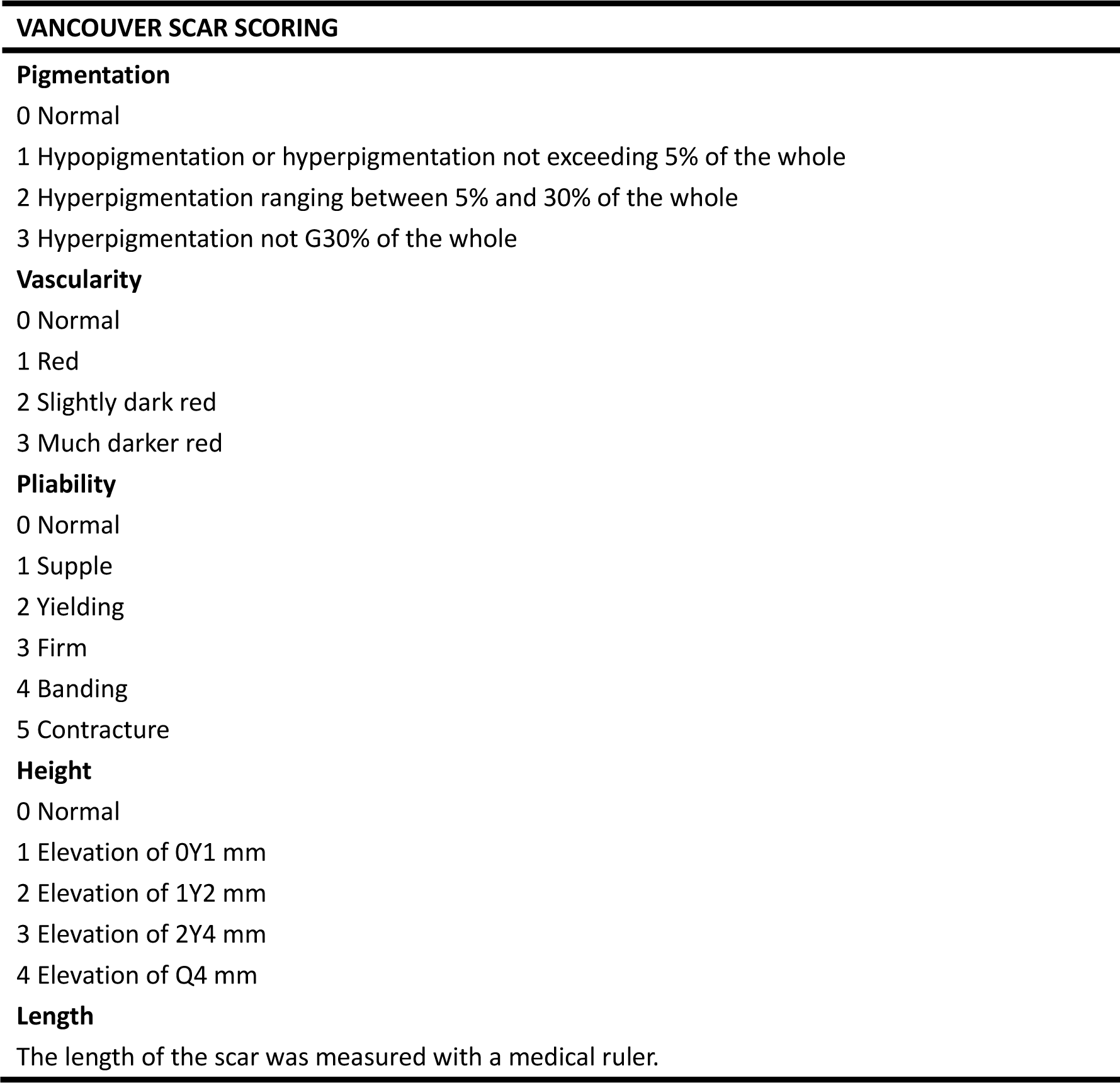
VANCOUVER SCAR SCORING(VVS)

The main adverse eactions include local pruritus, redness, swelling, and pain after treatment; a small number of patients can develop pain at the treatment site at the end of treatment; clinical treatment adverse reactions are classified into the following four levels, as shown in Table 3. In the event of an adverse reaction that is not tolerated by the patient, the follow-up physician can be contacted at any time to ask whether the drug needs to be discontinued or treated for the adverse reaction. The specific implementation of such measures is reflected in Table 4.

**Tables3.**
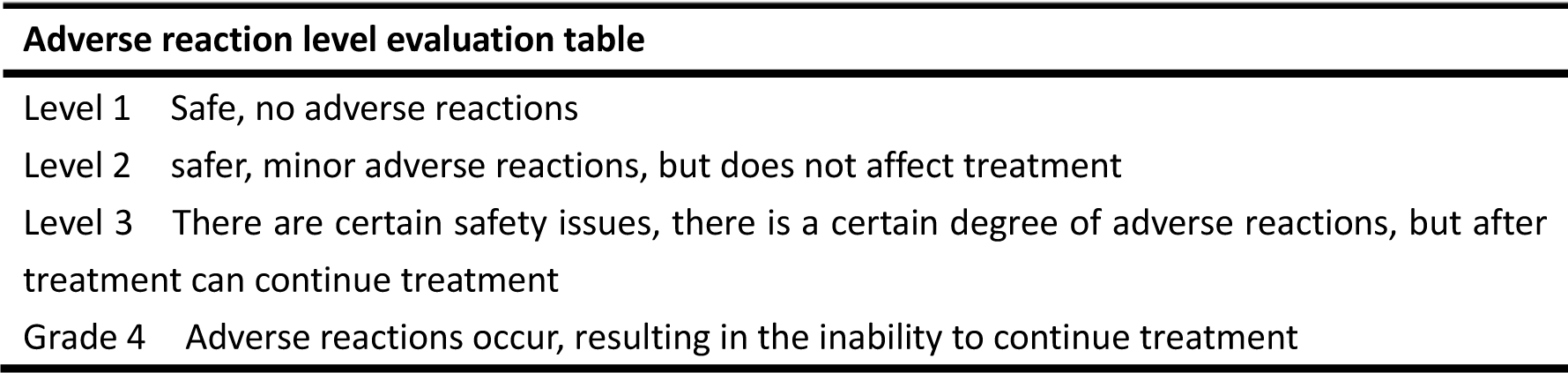
Adverse reaction level evaluation table.

**Table 4.**
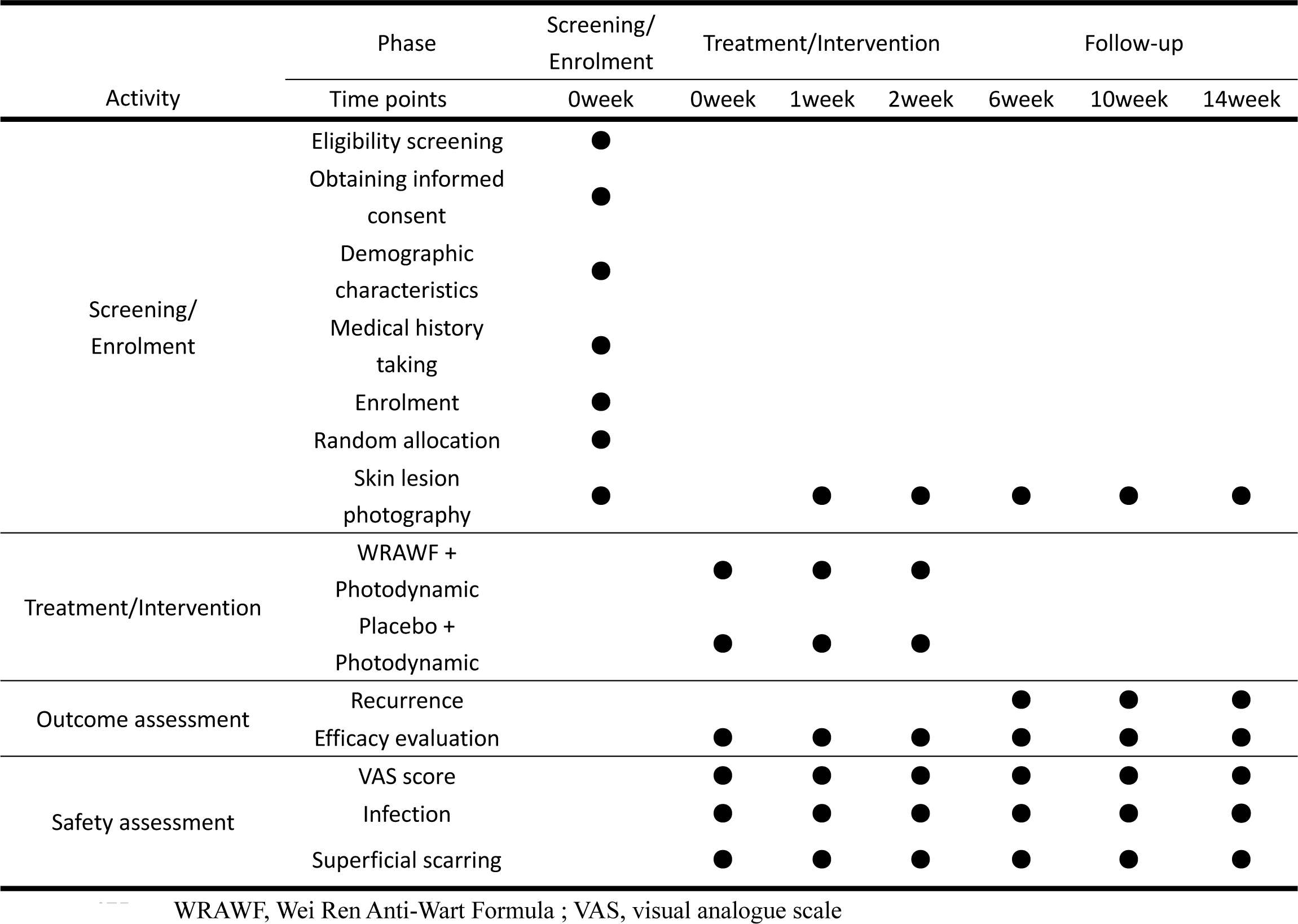
Schedule for enrollment, intervention, and assessment.

### Sample size

According to the literature,[14] the literature recurrence rate is 26.25%. According to the following formula, we assume that the recurrence rate in the combined treatment group is 8.7%, taking α=0.05 level and test efficacy of 0.8, then 1-β=1-0.2=0.8, two-sided test, using two sample rates to compare the sample content estimation formula. The rate for the test group was p_1_ and the rate for the control group was p_2_, yielding a sample content of 70 cases per group, assuming a 5% loss of patient follow-up; a total of 148 patients needed to be observed, 74 cases per group.

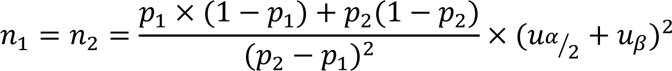

### Randomization and allocation

The trial was completely randomized and was conducted by the Data Management Center of Jiangsu Famaxon Medical Technology Co. The system will give a subject identification number, i.e., subject ID, as a unique identification number after enrollment. After obtaining the subject ID, the subjects were grouped by logging into the randomization center website, and the corresponding randomization number was obtained to enter the group.

### Treatment cycles

The study period was 15 weeks in total: photodynamic therapy (3 weeks) + follow-up period (12 weeks).

### Test drugs and blinding

The study drug and its placebo were manufactured, packaged, and provided by Sichuan Neo-green Pharmaceutical Technology Development Co., Ltd. The study drug randomization code was the unique identification code for the subjects. A double-blind design was used, and the randomization code table was created by the research randomiser program (https://www.randomizer.org/). Each participant’s corresponding PID will be given a unique random number and a corresponding drug packet or placebo packet with the same appearance, odor, and color. The project team will designate separate dispensers who will not be involved in other parts of that clinical trial. The monitors and outcome evaluators had to be blinded from the beginning to the end.

Unblinding regulations: This study provides for one unblinding. The unblinding is done after the blinded verification data is locked, and the staff member who keeps the blinded base will give it to the statistician for statistical analysis.

Emergency unblinding: When a serious adverse event occurs, or when a patient needs to be resuscitated and other circumstances necessitate knowledge of the drug used by the patient, the person in charge of the central trial will decide to unblind the patient in an emergency and open the corresponding emergency letter. Once the emergency letter with the corresponding number is opened, the case will be treated as a dislodged case.

### Drug combination

Any medications taken at study entry and during the three months prior to study, entry must be recorded on the Case Report Form (CRF). All medications used during the study are considered to be combined medications and must also be recorded on the CRF form. In particular, all permitted or unpermitted use of any measures for the treatment of psoriasis should be recorded. Combination medications should be recorded by type (trade name), dosage, indication, and duration of administration on the appropriate CRF page. The investigator decides whether the patient withdraws from the study.

I. Permissible combined treatments: certain drugs/other treatments that must be taken by patients with comorbid diseases or new comorbid diseases during treatment will be permissible, provided that the name of the drug (or other treatment names), dosage, number and duration of use, etc., are recorded in the study medical record for analysis and reporting at the time of summary.
II. Prohibited combined treatments: other internal medications, topical preparations, physiotherapy, etc. to treat the disease, including vitamin D3 derivatives, other herbal preparations, etc.

### Statistical analysis

Data management was performed using the data management platform designed by the Data Management Center of Jiangsu Famaxon Medical Technology Co., Ltd, which included data management, blinded audit, and data export were performed in this platform for data entry. The analyses will be performed using SAS statistical software and will include the actual number of subjects enrolled in both groups, disenrollment and exclusion cases, demographic and other baseline characteristics, adherence, efficacy analysis, and safety analysis.

Statistical analysis was performed using descriptive statistical analysis and comparative analysis between the two groups. Qualitative indicators were described by frequency tables, percentages, or composition ratios; quantitative indicators were described by means, standard deviations, or medians, lower quartiles (Q1), upper quartiles (Q3), minimums, and maximums. Qualitative data were tested by the chi-square test, Fisher’s exact probability method, Wilcoxon rank sum test, and CMH test. Quantitative data conformed to a normal distribution by t-test (chi-square test between groups, with 0.05 as the test level, and Satterthwaite’s method was used for corrected t-test when the variance was not equal), and Wilcoxon rank sum test and Wilcoxon signed rank sum test were used for non-normal distribution. Hypothesis testing was uniformly performed using a two-sided test, giving the test statistic and its corresponding P value, with P ≤ 0.05 as statistically significant and P ≤ 0.01 as highly statistically significant.

### Adverse events

I. Recording of adverse reactions or adverse events: ① The investigator should explain to the patients and ask them to respond truthfully to the changes in their condition after medication and avoid induced questions; ② While observing the efficacy, pay close attention to the observation of adverse reactions or unanticipated toxic side effects, analyze the causes, make judgments, and follow up the observation and records, and count the incidence of adverse reactions; ③ For the adverse reactions occurring during the study period, the symptoms, the degree, time of appearance, duration, treatment measures, and passage should be recorded on the case report form to evaluate its relevance to the test drug, and be recorded in detail by the investigator, signed and dated.
II. Reporting of adverse reactions or adverse events: ① When adverse reactions are found, the observing physician may decide whether to discontinue observation according to the condition, and cases of discontinuation due to adverse reactions should be followed up and investigated, with detailed records of treatment and results; ② If serious adverse events occur in the trial, the unit undertaking the clinical study must take immediate measures to protect the safety of the subjects and promptly inform the participating research units, and Ensure that all legal and regulatory requirements for reporting procedures are met.

### Data management and monitoring

The data management of this project was entrusted to the Data Management Center of Jiangsu Famaxon Medical Technology Co. This project uses the clinical data management platform, and the data manager constructs e-CRF according to the study protocol and case report form.

I. Data record In view of the situation that most of the outpatient medical records in China’s hospitals are brought by the patients themselves, to keep the first-hand data of the clinical trial intact, we design this trial-specific: “study medical record”. The study medical record is the source document of the clinical trial subjects, which should be kept in the hospital. The study medical record is the medical record of the outpatient subjects, and together with the inpatient medical record, it forms the medical record of the subjects.
II. Study case record requirements The investigator must write the study case record at the same time as the subject is seen and treated to ensure that data are recorded in a timely, complete, accurate, and truthful manner. Any substantiated corrections made to the study medical record may only be crossed out, with the corrected data noted next to it, signed and dated by the investigator, and not erased or overwritten on the original record. Original lab sheets for outpatient subjects are affixed to the study medical record and original lab sheets for inpatient subjects are affixed to the inpatient medical record. The results of laboratory tests for both outpatient and inpatient subjects must be completed in the “Physical and Chemical Findings Report Form” in the study medical record.
III. Review of medical records After each subject’s observation session, the investigator should submit the “study medical record”, “informed consent” and “patient medication record card” to the principal investigator of the unit within 3 working days. Within one week, they should be submitted to the project leader for review and deposited in the institutional data archive, and any problems found should be dealt with and recorded on time.
IV. Data Entry and Modification Data entry and management are the responsibility of the clinical coordinator and data manager, respectively. Each study center will be given a separate username and password before the start of the project, and a designated person from the study center will enter the raw data into the EDC via a secure network. Study center personnel will be allowed to make changes to the entered data during the entry process. The supervisor confirms that all data in the EDC is consistent with the original record. If there are errors or omissions, a challenge form is sent to the research center personnel to request changes to the data in the EDC. The data management department reviews the data recorded in the EDC by standard operating procedures. For queries in the data, the data manager will send a query to the research center personnel in the form of a query form, requesting the designated personnel of the research center to respond to the query and make the necessary changes to the data. All data changes and related operations shall be recorded with a trace.
V. Data Monitoring The number of monitors and the number of visits must meet the quality control requirements of the clinical trial. Supervisors shall review each study medical record and fill in the “Supervisor Review Page” one by one. During the monitoring process, they should check the completeness of the case records and the accuracy of the case report form, verify the trial data, check the compliance with the trial protocol and clinical trial management specifications, understand the progress of the enrolled subjects, and ensure the storage, distribution, and use of the trial products. The investigator and related personnel should assist the monitor during his or her visit and arrange an appropriate workspace for him or her.
VI. Database locking After a blind audit and confirmation that the database created is correct, a data management audit report is written by the data manager, and the data is locked by the principal investigator, sponsor, and statistical analyst. No further changes will be made to the locked data files.

## DISCUSSION

CA is a proliferative genital perianal lesion caused by infection with the human papilloma (HPV) virus. Its incidence is three times higher than that of genital herpes, and most patients are young people aged 18-35.[15,16] Thev infection of warts not only has physical and psychological effects but the pathogen HPV is also carcinogenic in addition to causing warty growths.[17] Evidence suggests that HPV infection is closely associated with the development of cervical cancer.[18]

The typical CA is a small, reddish papule that gradually increases in size and has an uneven surface, and then further proliferates into a wart-like protrusion and spreads to the periphery. The most important thing is that you can feel the pain of intercourse, and occasionally the infection may be oozing, ulcerated, or bleeding.

The principles of CA treatment are to remove as many warts as possible and to reduce recurrence. Although there are several treatments for warts, there is a risk of recurrence with any of them.[19] There is no definitive answer as to which method is the best for treating warts.

In Traditional Chinese Medicine (TCM), it is mainly contact with evil toxins, and damp heat, which is caused by the accumulation of damp heat toxins on the pudendal skin. The TCM syndrome types of this disease are mainly divided into two types: damp-heat toxin accumulation syndrome and dampness and toxicity gather in the lower, while CA is mainly localized, so external treatment is often used for treatment. The effect of drugs can penetrate the skin directly to the affected part.[20] In China, the compound external preparation mainly composed of Brucea javanica, Sophora flavescens, Honeysuckle, Folium Isatidis, diffusa diffusa, honeydew, fructus cnidii, and other traditional Chinese medicines has been used for many years. Studies have shown that it can inhibit and kill HPV through cytotoxicity.[21,22] There are also external preparations of cantharidin, which are effective in the removal of the wart body and the prevention of recurrence. However, due to the difficulty of standardization and the limitations of the research methods adopted, there is a lack of relevant high-quality evidence-based medical evidence.

In this trial, we will use granular formulations to minimize bias associated with the use of herbs from different geographic regions, different species, and preserved under different storage conditions. In this study, it refers to clarify the superior aspects of Chinese medicine in the treatment of CA and to obtain high-level evidence-based medical evidence to provide a basis for the formation of standardized treatment protocols.

### Ethics and dissemination

This trial has been ethically approved by the Ethics Committee of Shanghai Yueyang Hospital of Integrative Medicine (approval number:2023-023) All participants will be enrolled only after signing the informed consent form. No clinical data or biological samples will be collected from those who do not participate. The trial will comply with national laws, clinical practice guidelines, and the Declaration of Helsinki as revised in 2013. Results will be published through meeting reports and publicly available documents.

### Trial status

The trial phase begins in JAN 2024 at Shanghai Yueyang Hospital for Integrative Medicine and is currently being prepared for patient recruitment.

### Contributors

**SYY** drafted the manuscript. LH participated in the design of the study, and YFW coordinated the study. WXQ will generate the allocation sequence, DJG will enrol participants, and XQW and FLL will assign participants to interventions. All authors read and approved the final manuscript.

### Patient consent Patient

recruitment has not started.

### Ethics approval

Institutional Ethics Committee of Shanghai Yueyang Integrated Medicine Hospital.

## Data Availability

All data produced in the present study are available upon reasonable request to the authors
All data produced in the present work are contained in the manuscript

